# Deaths averted by vaccination due to reduced transmission can exceed those from direct protection of vaccinated individuals for SARS-CoV-2

**DOI:** 10.1101/2021.10.25.21265500

**Authors:** Billy J. Gardner, A. Marm Kilpatrick

## Abstract

Vaccination programs often focus on direct protection of vaccinated individuals against disease and discount reductions in transmission, which can result in preventable disease and death. Initial clinical trials for most COVID-19 vaccines only measured direct protection, and dosing and vaccine selection decisions have, so far, ignored effects on transmission. Here we provide a novel empirical framework for incorporating effects of COVID-19 vaccination on transmission in a continuous dynamic immune landscape. We quantified relationships between neutralizing antibody titers and vaccine effectiveness for both susceptibility (VE_S_) and infectiousness (VE_I_) and quantified changes in VE with waning and boosting of immunity. We used these relationships to quantify the impact that additional doses of mRNA vaccines (BNT162b2 and mRNA-1273) could have had in reducing transmission and deaths caused by the deadliest SARS-CoV-2 variant, Delta, in Autumn 2021. Neutralizing antibodies waned 8-fold in 2021 over the six months following initial vaccination with mRNA vaccines, which reduced VE_S_ 33-38% (from 75-81% to 47-54%) and VE_I_ 62-65% (from 47-57% to 16-22%) against the Delta variant. Third doses increased neutralizing antibody titers 13-26-fold, which more than restored VE and reduced the relative risk of transmission 7-10-fold. Administering third doses by September 1, 2021, could have reduced the effective reproductive number R_t_ by 19%, stopped surges in transmission in many populations, and averted an estimated 113,000 deaths in the United States, which is 2.6-12.4 fold higher than direct effects in vaccinated individuals. Vaccination programs that incorporate effects on transmission in trial design, vaccination frequency, and vaccine choice are needed to address current and future public health challenges.

## Introduction

Vaccination is the most effective tool for reducing disease, hospitalizations, and deaths for many infectious diseases ^1–5^. Core components of vaccination programs include how to allocate vaccine doses within and between individuals, when to recommend new doses and, when multiple vaccines are available, which vaccines to recommend. In making these decisions and developing new vaccines, public health agencies often focus on the direct impact of vaccines in reducing disease in vaccinated individuals ^6,7^. For example, the United States Food and Drug Administration guidelines for randomized control trials for COVID-19 vaccines did not include any criteria regarding protection against infection or reductions in infectiousness ^8^, and, as a result, these weren’t measured in the US trials for mRNA vaccines made by Pfizer-BioNTech and Moderna. Similarly, vaccines are often only recommended for subsets of the population at high risk disease severity ^9^, despite the fact that in many cases, such as pneumococcal disease, vaccines have been shown to reduce transmission ^10^.

Many vaccines can have population-wide benefits by reducing transmission either by reducing susceptibility or reducing infectiousness (i.e. transmission from breakthrough infections), or both, which indirectly protects additional individuals ^11^. In these cases, vaccinating individuals who are not at risk of severe disease can reduce severe disease through reductions in transmission ^12^. Deploying vaccines to reduce transmission can increase health equity by reducing infection in individuals that lack access to health care, including vaccination, which is a key global challenge ^13^. However, determining whether the benefits of deploying vaccines to reduce transmission is warranted requires estimating vaccine effectiveness (VE) for transmission and the potential number of deaths averted for a specific population, which, in turn, depends on the dynamic levels of population immunity for the pathogen in question.

The emergence of the Delta variant (B.1.617.2) of SARS-CoV-2 in 2021 illustrates the potential benefit of using vaccination to reduce transmission. The Delta variant caused a surge of deaths globally, even in populations with high initial two-dose vaccination coverage ^14^. This was due, in part, to the higher infectiousness and severity of this virus variant ^15,16^, partial immune evasion ^17^, and, critically, waning vaccine immunity. Waning immunity was evident in both levels of neutralizing antibodies and studies of vaccine effectiveness, including for severe disease in older individuals ^18,19^. Following the Delta variant surge in autumn 2021, third vaccine doses were initially offered to older ages and at risk individuals, but not to healthy younger individuals because VE for severe disease in these age groups remained relatively high ^20^. However, the potential impacts of offering third doses to all individuals to reduce transmission was not considered because of a lack of information on how much it could reduce transmission_21._

Here we estimate the potential impact of third doses of mRNA vaccines on population-level transmission of the Delta variant in late 2021 and the number of deaths that could have been averted. We first estimated how vaccine effectiveness for both susceptibility (VE_S_) and infectiousness (VE_I_) change over time due to waning and boosting of immunity. Previous work has demonstrated strong correlations between a surrogate measure of protection, neutralizing antibody titers, and VE for symptomatic and severe disease ^22–27^, and one study used these relationships to examine the impacts of vaccination for averting disease and deaths^26^. Here, we extend this approach by developing the first relationships, for any pathogen, between neutralizing antibody titers and VE_S_ and VE_I_, which are required to estimate the impact of vaccination on transmission, as well as vaccine effectiveness for death, VE_D_. We then quantified relationships between neutralizing antibody titers and time since vaccination, and following boosting with a third dose of mRNA vaccines. We integrated these two relationships to estimate the potential impacts of deploying third vaccine doses to varying fractions of doubly vaccinated individuals on the reproductive number of the virus, *R_t_,* under multiple scenarios. Finally, we estimated the number of Delta infections and deaths that could have been averted in the United States in Autumn 2021 by deploying third doses, and compared the effects of reducing transmission to the deaths that may have been averted due to the direct effects of third doses on protection against death.

## Results

We found very strong relationships between variant- and vaccine-neutralizing antibody titer ratios (NATR_tot_) and VE using 23 estimates of VE_S_ (susceptibility) and 14 estimates of VE_I_ (infectiousness) across three variants and four vaccines (Figure 1A,B; Table S1). VE_S_ was higher than VE_I_ across the range of NATR_tot_ observed, but the slope of VE_I_ was higher (VE increased more rapidly as NATR_tot_ values increased) (Figure 1; Table S1).

**Fig. 1.**
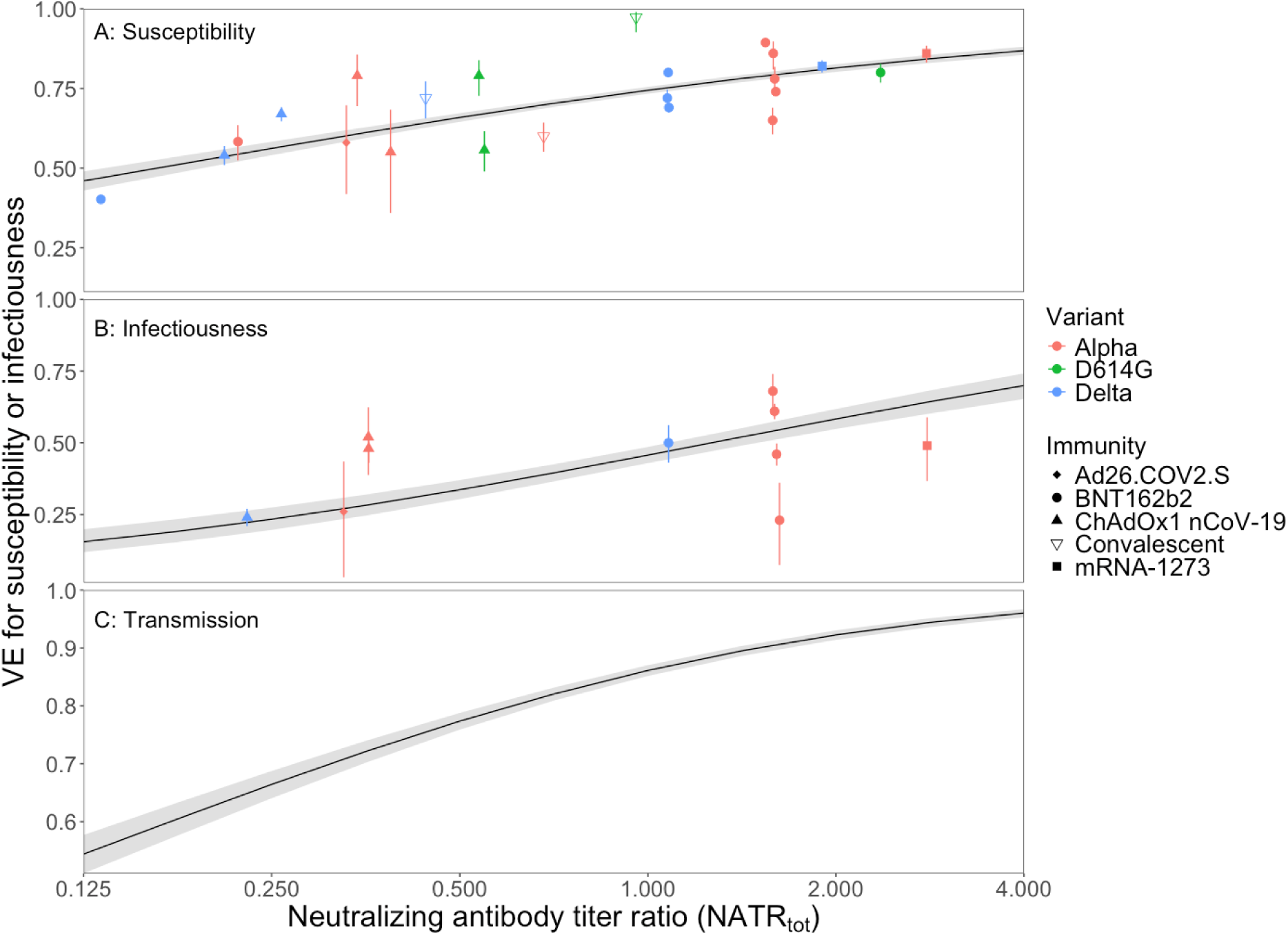
Vaccine Effectiveness (VE) and variant- and vaccine-specific neutralizing antibody titer ratios (NATR_tot_). VE for (A) susceptibility, VE_S_, (B) infectiousness, VE_I_, and (C) transmission VE_T_ plotted against NATR_tot_. Each point (± 1 SE) represents a single empirical estimate of VE for a single vaccine (or infection) & virus variant. Points are jittered slightly along the x-axis to facilitate presentation. Black lines and ribbons in (A) and (B) show the fitted model and 95% CIs. The line and 95% CI ribbon in (C) is estimated from lines in (A) and (B): VE_T_ = 1-(1-VE_S_)*(1-VE_I_); see Methods for details.

In the eight months after vaccination, neutralizing antibody titers waned approximately 8-fold for vaccine-derived immunity for both BNT162b2 and mRNA-1273 vaccines and 3-fold for infection-derived immunity, with most waning occurring in the first 3-5 months (Figure 2A, B; Table S2). Initial neutralizing antibody titers were highest for vaccination with mRNA-1273, followed by BNT162b2, and lowest following infection (Figure 2A; Table S2). Waning rates were faster for the BNT162b2 vaccine than the mRNA-1273 vaccine and infection-derived immunity (Figure 2B; Table S2).

**Figure 2.**
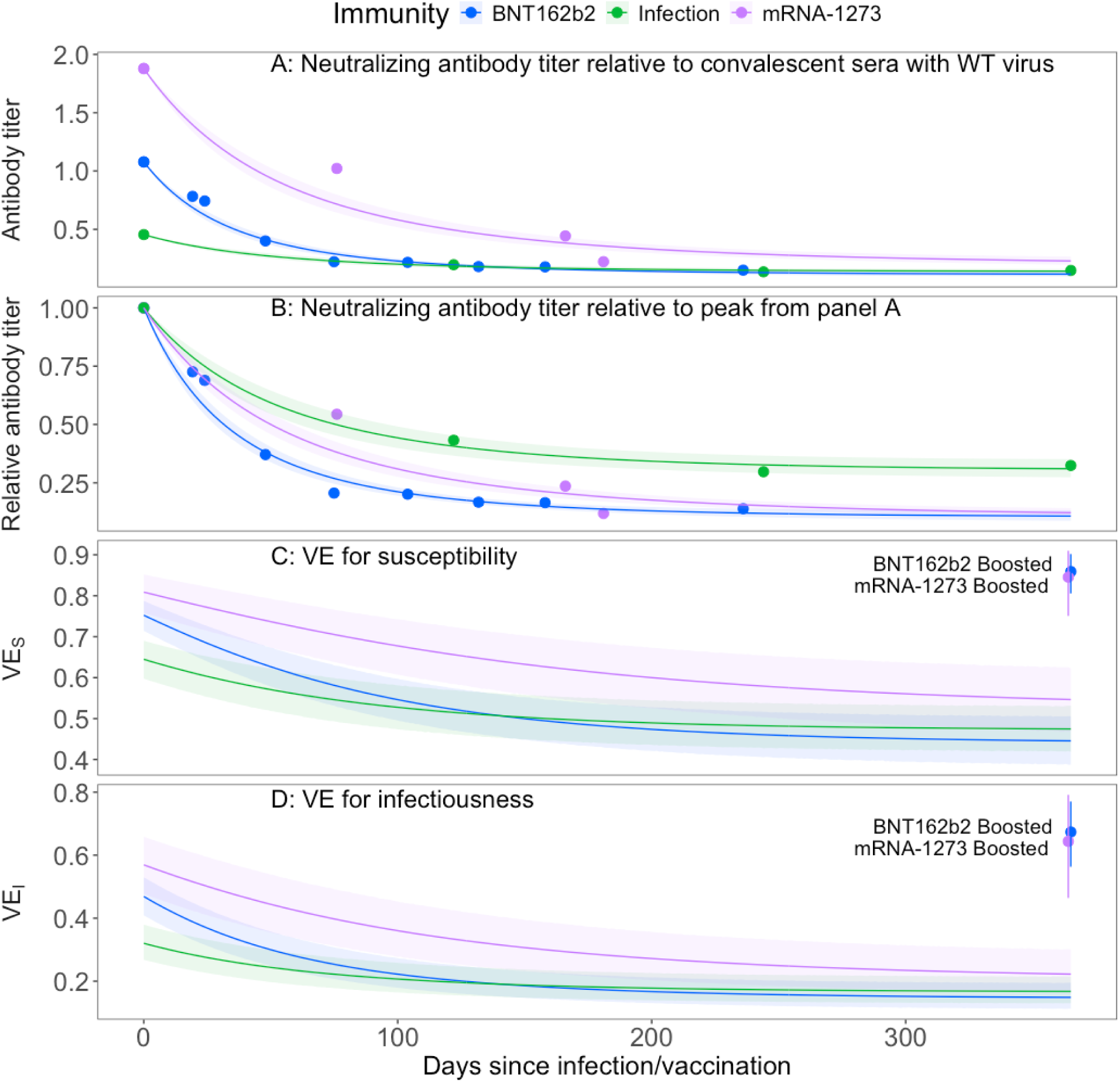
Waning of neutralizing antibody titers and VE for susceptibility (VE_S_) and infectiousness (VE_I_) over time and following boosting with a third vaccine dose. Lines show fitted models and ribbons show ± 1 SE (Table S1). A) Neutralizing antibody titers relative to convalescent sera with wild-type (WT) virus, adjusted for the 2.2-fold lower neutralization titers with the Delta variant relative to wild-type virus. B) The same data as in panel A but rescaled relative to the initial peak value (i.e. all lines start at a value of 1) to show relative rates of waning and levels of stabilization (Table S1). C) VE for susceptibility (VE_S_) over time using the patterns of waning in Panel A and the relationships between antibody titers and VE for susceptibility (Figure 1A). D) VE for infectiousness (VE_I_) over time using the patterns of waning in Panel A and the relationships between antibody titers and VE for infectiousness (Figure 1B). Points in the upper righthand corner of panels C and D, indicated by the black arrows, show the effect of third doses in boosting VE for the BNT162b2 and mRNA-1273 vaccines.

We used the relationships between neutralizing antibody titers and VE (Figure 1) and patterns of waning of antibody titers (Figure 2A, B) to estimate the waning of VE_S_ and VE_I_ over time (Figure 2C, D). As expected from levels of antibody titers, VE_S_ and VE_I_ were highest for two-dose vaccination with mRNA-1273, and BNT162b2 vaccines, followed by infection-derived immunity (Figure 2C, D). The relatively fast waning rate of neutralizing antibodies for the BNT162b2 vaccine suggests that VE for this vaccine was initially much higher than infection-derived immunity, but the two were similar after 3-4 months (Figure 2C, D). VE for the mRNA-1273 vaccine remained higher than both the BNT162b2 vaccine and previous infection as protection waned (Figure 2C, D).

A third dose of the BNT162b2 vaccine boosted antibody titers 26-fold relative to levels after 8 months of waning, or 25.9/8.1 = 3.2-fold higher than one week after dose two ^28^. A third dose of the mRNA-1273 vaccine boosted antibody titers 13-fold relative to 8-month waned levels, or 1.5-fold higher than shortly after the second dose. The fitted relationship between VE_S_ and neutralizing antibody titer ratios (Figure 1) suggested that third doses of the mRNA vaccines would increase VE_S_ 1.6-1.8-fold from waned values of 47-54% to boosted values of 85-86% and would boost VE_I_ 3-4-fold from 16-22% to 64-67% (Table S3; Figure 2C,D; compare blue and purple points in upper right of panels to lines of the same color below). Combined, boosting waned individuals with a third vaccine dose increased VE for transmission (VE_T_ = 1-(1-VE_S_)*(1-VE_I_)) from 55.4% to 95.4% for BNT162b2 and from 64.3% to 94.5% for mRNA-1273. In doing so, boosting reduced the risk of transmission (1-VE_T_) by ∼10-fold or 89.7% for BNT162b2 and ∼7-fold or 84.5% for mRNA-1273 vaccinated individuals compared to waned immunity. In comparison, VE against death (VE_D_) waned from 96.5% to 92.1% for BNT162b2 and from 97.2% to 93.5% for mRNA-1273, with boosting increasing protection to 97.8% and 97.6%.

Next, we examined the impacts of third doses on population-level transmission in the United States via reductions in the reproductive rate of the pathogen, R_t_. On September 1, 2021, the average time since vaccination for the 54.6% of the population that had been vaccinated was 4 months (111.4 days) (Figure S1), resulting in average antibody titers waning to less than 22% of initial peak values. If all doubly vaccinated individuals in the USA (54.6% of the population) had received a third dose of an mRNA vaccine on 25 August 2021 (becoming maximally effective on 1 September 2021^29^) this would have increased VE_s_ and VE_I_ as described above and reduced *R_t_* by 19.3% (95% CI: 15.5% to 22.3%) from 1.19 to 0.96 (Figure 3D). This would have stopped the strong surge in cases, hospitalizations, and deaths that occurred over the next few months, assuming contact rates and other behaviors didn’t change following boosting (see Methods for further details).

**Figure 3.**
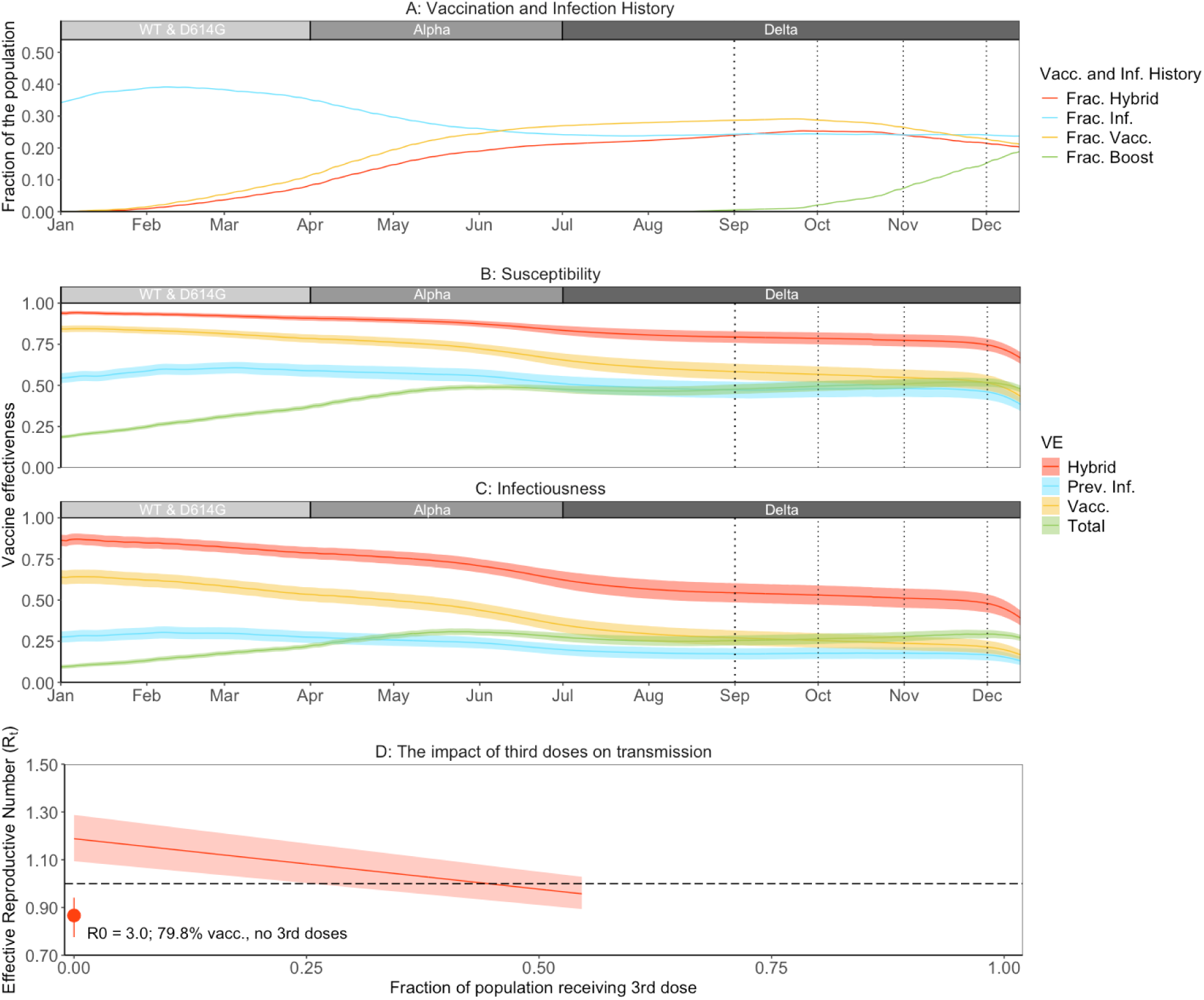
United States population vaccination and infection history over time, population-level vaccine effectiveness against susceptibility (VE_S_) and infectiousness (VE_I_) over time, and the relationship between the pathogen effective reproductive number, *R_t_* and third dose coverage, as of 1 September, 2021. A) Fraction of the US population that had received their primary vaccination series but had not been infected (Frac. Vacc.), had been infected but not vaccinated (Frac. Inf.), had been both vaccinated and infected (Frac. Hybrid), or had received their third-dose booster (Frac. Boost). B) and C) Lines ± 1 SE show population-level VE against susceptibility (B) and infectiousness (C) over time for three subpopulations (vaccinated with their primary series, previously infected, and both vaccinated and previously infected [hybrid]) and the entire population in the US (“Total”) which is a mixture of the five subpopulations, as shown in panel (A). Rectangles above panels (A-C) show which variant was dominant during the period (frequency ≥ 50%). D) Line ± 1 SE shows R_t_ plotted against the fraction of vaccinated individuals receiving third dose boosters for the United States with R_0_=3.0, 54.6% vaccinated, and 58.4% previously infected as of September 1, 2021 (see Methods for additional details). The single red point shows the impact of using all third doses from the right end of the red line to doubly-vaccinate unvaccinated individuals, which would bring the vaccination coverage from 54.6% to 81.8%. The dashed horizontal line shows the threshold reproductive number R_t_ = 1, separating growing from shrinking epidemics.

We explored the possibility of using the same number of doses to doubly vaccinate unvaccinated individuals, assuming they were willing to be vaccinated, rather than boost previously vaccinated individuals. This would have been even more effective in reducing transmission. R_t_ would have decrease from 1.19 to 0.87 versus 1.19 to 0.96 for using doses for boosting vaccinated individuals (Figure 3D, compare right end of red line to red point labeled “R_0_ = 3.0, 81.8% vacc., no 3^rd^ doses” on left side). We also explored several other scenarios with varying vaccination and infection rates and contact rates which matched characteristics of other populations. These analyses showed that the percent reduction in R_t_ was driven primarily by the fraction of the population vaccinated, which, in turn, determined the potential boosting fraction (Supplemental Results Text, Figure S2).

We then calculated the number of deaths that could have been averted through reduced transmission by administering third doses. Between June 2021, when the Delta variant emerged, and January 2022, when Omicron fully displaced the Delta variant, the Delta variant caused an estimated 273,801 deaths ^14^. The average number of estimated daily cases sometimes exceeded 200,000 (Figure S3), and reported deaths sometimes exceeded 2,000/day (Figure 4). However, during this period the estimated R_t_ for the US population mostly fluctuated between 0.9 and 1.1 (Figure S3), suggesting that many thousands of deaths could have been prevented if R_t_ could have been reduced, even marginally. As just described, boosting the 54.6% of the population that had been doubly vaccinated with a third dose by 1 September, 2021 would have reduced R_t_ by 19.3% (Figure 3D, red line). This would have reduced the number of Delta deaths by approximately 113,000 (95% CI: 105,500 to 117,600) (Figure 4E, blue shaded region) by averting 37,700,000 infections (95% CI: 35,200,000 – 39,300,000) (Figure 4A, orange shaded region). This corresponds to 637.8 (95% CI: 595.5 to 663.9) deaths averted per million vaccine doses administered due to effects on transmission alone.

**Figure 4.**
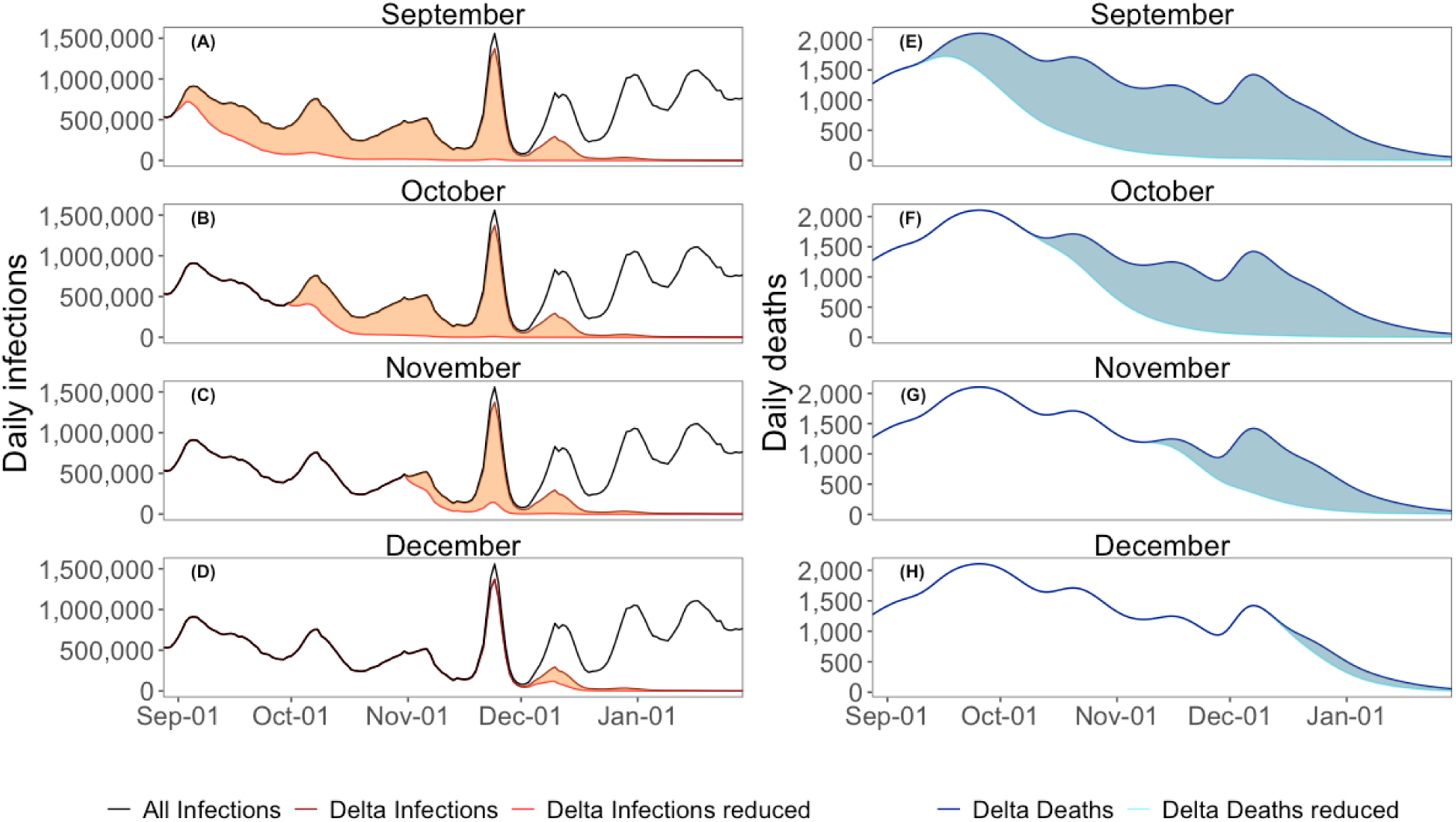
Estimated daily infections and reported or estimated deaths with and without third dose boosting plotted against time in the United States when the Delta SARS-CoV-2 variant was dominant. Black lines in the left panels (A-D) show the estimated number of infections each day, calculated by deconvolution using the observed daily deaths (E-H; dark blue lines in right panels) and the delays between infection and death, and using and an infection fatality ratio (IFR) of 0.003 ^30^. Deaths from the Delta variant were estimated using reported deaths and variant fractions based on sequencing data (see Methods). Red lines in left panels show the estimated number of infections with third dose boosting, and light blue lines show the number of deaths resulting from these infections with the timing estimated by reconvolution given the same distributions. The shaded regions show the averted infections (orange) and deaths (blue) if all eligible vaccinees received third doses that took full effect on Sept. 1, Oct. 1, Nov. 1, or Dec. 1 as indicated on the panel title with the reduction in R_t_ for these dates shown in Figure S2.

We compared these indirect effects on transmission to the direct effects of giving third-dose boosters, as well as the direct effects of using those same doses to give two doses to the unvaccinated. The direct impacts of third-dose boosters would have reduced deaths by only 9,200 (95% CI: 8,400 to 9,800) (Table S4), and the direct impacts of distributing those same doses to unvaccinated individuals would have reduced deaths by 44,900 (95% CI: 44,400 to 44,200) (Table S4). Both of these estimates are far lower than the deaths averted due to indirect effects described above (113,000).

Boosting populations later (i.e. administering vaccines so VE would reached maximum effectiveness Oct.. 1, Nov. 1 or Dec. 1) would have resulted in greater reductions in R_t_ as immunity in the vaccinated group continued to wane, but would have averted fewer deaths caused by the Delta variant. Deploying third doses that resulted in peak VE on October 1, November 1, or December 1 would have reduced R_t_ by 30.3%, 31.4%, and 30.3%, respectively (Figure S2), but would avert only 74,900 (95% CI: 70,300 – 76,000), 36,500 (95% CI: 33,700 – 38,100) and 6,000 (95% CI: 5,300 – 6,500) Delta deaths (Figure 4; Table S5).

## Discussion

The indirect effects of vaccination in reducing disease by reducing transmission (via reduced susceptibility or infectiousness) were rarely considered at any stage of COVID-19 vaccination programs, including initial trial design, initial vaccination, and subsequent boosting. We found that boosting population immunity with third vaccine doses could have reduced transmission of the most deadly SARS-CoV-2 variant, Delta, stopped surges, and averted a huge number of deaths in many populations, including more than 100,000 in the US alone. This estimate of 638 averted deaths per million vaccine doses is similar to or higher than the direct effects of COVID-19 vaccination in other studies (93 to 836 deaths per million doses administered ^31–34)^. Unfortunately, many countries initially offered third doses only to protect older and at-risk individuals and waited until later to offer third doses to the general population^20^. Possibly as a result, third dose uptake by the general population in most countries was low. It may have been higher if third dose boosters were immediately and strongly recommended, or mandated for the whole population ^35^.

The approach we developed here provides an empirical framework for estimating the effect of vaccination on transmission in a dynamic immune landscape. The key components needed for this approach are the temporal dynamics of a surrogate of protection (in our case, neutralizing antibody titers) and relationships between neutralizing antibody titers and the two components of transmission: susceptibility and infectiousness. Previous studies on COVID-19 have shown that neutralizing antibody titers were strongly correlated with VE for symptomatic disease and severe disease ^22–24,26,27^. Here, we significantly extended this approach by showing that antibody titers were also strongly correlated with VE_S_ and VE_I_. This advance is critical for building accurate mathematical models of SARS-CoV-2 transmission as immunity wanes and is boosted with additional vaccine doses or infection. Previous models have assumed VE for transmission was the same as VE for symptomatic disease^26^, which overestimates VE for susceptibility (the relative risk of infection) and underestimates VE for transmission (Figure S4). A similar empirical framework for estimating the effect of vaccination on transmission could be developed for other pathogens, such as influenza virus, if a suitable surrogate of protection^36^ and multiple estimates of vaccine efficacy for susceptibility and infectiousness in population with different levels of immunity could be obtained.

We found that VE_S_ and VE_I_ declined rapidly in the first 3-5 months post-vaccination in BNT162b2 recipients and plateaued after ∼6 months, with slower, waning in mRNA-1273 recipients, which may or may not be related to long lasting plasma cells ^37,38^. These rates of waning suggest that vaccinating individuals multiple times per year would have substantially increased VE for transmission against the Delta variant, and may increase VE for transmission for the currently circulating SARS-CoV-2 variants as well. Given the summer surges of COVID-19 hospitalizations that now appear to be a repeatable phenomenon, recommending two vaccine doses per year for COVID-19, especially in sensitive settings such as skilled nursing facilities, could provide substantial benefits.

We found that third vaccine doses could have reduced deaths caused by the Delta variant dramatically, but the impact depended on the timing of the administration of the doses. Rolling out third doses to reach peak VE by September 1 would have averted an additional 39,100 Delta deaths in the US compared to waiting until October 1, emphasizing the timeliness of vaccinations. Two related factors made a September 1 rollout especially impactful. First, infections were near their peak in September. Second, R_t_ was below 1.03 from September 1 until late November when the Delta variant faded in importance. The potential reduction in R_t_ we estimated was possible with third doses (19.3%) was sufficient to reduce R_t_ well below 1 which would have driven Delta infections and deaths down rapidly (Figure 4). Administering third doses later would have averted fewer deaths caused by the Delta variant, but may have averted additional infections and deaths caused by the Omicron variant. Additionally, we found that the deaths that could be prevented due to the impact of third dose boosters on transmission were substantially higher than those that would be averted due to direct protection alone.

Our study has several limitations. First, our analyses use population averages for estimates of VE_S_ and VE_I_ and do not incorporate age-specific variation among individuals. Age-specific patterns of contact, disease severity, and immunity can play a key role in shaping epidemics ^39^. We also assumed well-mixed populations in calculating reductions in the reproductive number R_t_. A targeted vaccination approach could be even more effective than that reported here if highly connected individuals could be targeted for third doses ^40^. Third, our calculations for the reductions in R_t_ with third doses were based on neutralizing antibody titers one month after the administration of a third dose. Neutralizing antibody titers also decay following third doses, but the decay rate is much slower than after second doses ^41^, suggesting that substantial reductions in R_t_ from third doses would have persisted for many months, as we assumed here. Fourth, our analyses were focused on population characteristics in the US in late 2021 and vaccine effectiveness against the Delta variant because data were available for quantifying immunity in populations at this time. The impact of vaccine doses on current transmission of SARS-CoV-2 is more difficult to quantify with currently available data, due to a more complex immune landscape, with most people having been infected several times with multiple virus variants and vaccinated with multiple vaccine types. In addition, the variant composition of SARS-CoV-2 viruses is sometimes diverse and sometimes changes frequently. Determining the current benefit of vaccination for reducing transmission would require estimates of the distribution of neutralizing antibody titers against currently circulating viruses in target populations, and measuring increases in antibody titers following vaccination. Finally, quantifying the impact of vaccination in reducing deaths due to reduced transmission does not take into account additional benefits of reduced infections including reduced absenteeism from work and school and reduced cases of long COVID. Taken together the total benefits of reduced transmission may often outweigh the costs of administering additional vaccine doses in many populations.

In summary, we showed that the indirect benefits of vaccination in reducing transmission can lead to a huge number of deaths averted. In addition, reductions in transmission increase health equity by reducing infection in individuals that lack access to vaccination. Quantifying vaccine impacts on transmission should be an integral part of developing new vaccines, and estimates of reductions in transmission should be used for planning vaccination strategies, including vaccine selection, targeting, and vaccination schedules.

## Methods

### Relative neutralizing antibody titers by variant, vaccine dose and with waning

We collected data on neutralizing antibody titers from the literature and estimated the total neutralizing antibody titer ratio (NATR_tot_) for each vaccine, virus variant, and immune status ^24^. NATR_tot_ is the ratio of neutralizing antibody titer for a given vaccine, variant and immune status, relative to the neutralizing antibody titer of convalescent sera after infection with the original wild-type virus ^24,27^. For example, two weeks after a second dose of the BNT162b2 vaccine NATR_tot_ for the Delta variant is 1.08, because vaccination with BNT162b2 results in neutralizing antibody titers 2.37-fold higher than infection ^24,27^, and neutralizing antibody titers against the Delta variant are 2.20-fold lower than against wild-type virus; 2.37*(1/2.2) = 1.08 ^24^. We also collected data from studies measuring neutralizing antibody titers over time in sets of individuals following infection or vaccination to quantify waning of neutralizing antibody titers, as well as increases in antibody titers following boosting with third doses^28,42,43^. We then fit models to the decay in antibody titers over time following vaccination with two mRNA vaccines (BNT162b2 and mRNA-1273), and infection (Table S1). We assumed antibody titers from hybrid immunity (infection followed by vaccination) waned at the same rate as following BNT 162b2 vaccination based on a study that measured both ^43^.

### VE for susceptibility, infectiousness, and death

We collected VE estimates for susceptibility, infectiousness, and death for SARS-CoV-2 from the literature, including a systematic living review ^44^, and categorized each study by vaccine, time since vaccination, and variant type (wild-type/D614G, Alpha, Delta). We excluded estimates where the virus variant could not be determined. For estimates of VE_S_, we only included studies where the endpoint was “all infections” (often measured using frequent testing or serology) and excluded studies where the endpoint was “any infection” because “any infection” studies do not capture all infections and include an unknown fraction of asymptomatic infections. Studies of VE_I_, or transmission given infection, usually compared secondary attack rates in households by vaccination status of the index case.

To fit a relationship between VE and NATR, we needed estimates of the number of infections in the vaccine and control groups. However, most studies did not report the number of infections; however, all reported a VE and 95% CI. We estimated the effective number of infected individuals in the control group (I_c_) and the effective number of infected individuals in the vaccine group (I_v_) for each study by determining the number of each needed to match the mean and 95% CI given in a study. We held the effective number of individuals in the control group (N_c_) and vaccine group (N_v_) constant at 1,000,000 because the 95% CI was invariant to variation in these values for observed incidence values. We used a maximum value of 1000 infections in the control group (I_c_) to reduce the undue leverage of some very large studies ^45^.

### Relationships between VE and neutralizing antibody titers by vaccine and variant

We modeled the relationship between VE and total neutralizing antibody titer ratios (NATR_tot_) using ^24^:

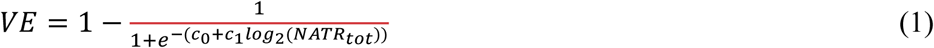

We estimated the coefficients c_0_ and c_1_ separately for the three endpoints, susceptibility, infectiousness, and death, by maximizing the likelihood of observing the data underlying the VE estimates: the number of infections in the control group, I_c_, and vaccine group, I_v_, for each study. The likelihood was the product of two binomial distributions with N_c_ and N_v_ individuals:

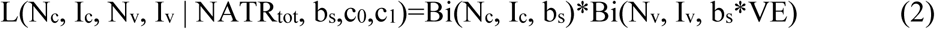

Here b_s_ is the baseline risk for the study period, which is the fraction of control individuals infected during the study. We maximized the likelihood across all studies which was the product of the likelihoods of the individual studies. We used the fitted relationships for the two endpoints, susceptibility and infectiousness, between VE and neutralizing antibody titer ratios, NATR_tot_ (eq 1) to estimate VE for populations based on the estimated NATR_tot_ given the timing of vaccination and infection and estimated rates of waning as described above. Finally we calculated the VE for transmission, VE_T_ = 1-(1-VE_S_)*(1-VE_I_). We calculated 95% CIs for VE_T_ using independent bootstrap samples from the VE_S_ and VE_I_ estimates and 95% CIs.

### Vaccine effectiveness over time

We estimated VE_S_ and VE_I_ in the US for each day from January 1, 2021 to December 13, 2021 for three subpopulations: previously infected and unvaccinated, twice vaccinated and no previous infection, and twice vaccinated and previously infected (hybrid immunity), the timing of prior infections and vaccinations (Figure S1), and the relationships between neutralizing antibody titers and VE_S_ and VE_I_. We estimated the timing of infections using reported deaths ^46^ and deconvolution, using the distributions for the delays between infection and death (i.e. infection to shedding, shedding to symptom onset, symptom onset to hospitalization, and hospitalization to death) (Table S6) using the deconvolve package in R ^47^. For each day, we estimated VE_S_ and VE_I_ in each subpopulation against each circulating variant based on the timing of infections and vaccinations, accounting for the waning that occurred in each subpopulation on that date using the fitted model for the relationship between neutralizing antibodies and time since infection and vaccination. We used sequencing data from GISAID ^30,48^ (offset by 9 days to account for the average delay between infection and reporting date) in the US to get a variant-averaged estimate of VE_S_ and VE_I_ in each subpopulation for each day using a weighted mean where weights were the observed variant frequency each day. We then estimated a population-wide VE for each day using estimates for the fraction of the population in five subpopulations: fully susceptible, third-dose boosted, and the three subpopulations described above.

### The impact of third doses on the reproductive number, R_t_

We used patterns of waning and boosting of neutralizing antibody titers to estimate the impact of providing additional doses of mRNA vaccines to increasing fractions of vaccinated individuals in the USA and other populations on the reproductive number, *R_t_*, of the Delta variant of SARS-CoV-2, at four time points in autumn 2021; Sept 1, Oct 1, Nov 1, and Dec 1 (Table S7). *R_t_* is the average number of secondary cases that each case infects and is equal to the basic reproductive number for a pathogen in a fully susceptible population, *R_0_*, multiplied by the fraction of the population that is susceptible (i.e. not immune from vaccination or previous infection).

We examined the effect of boosting vaccinated individuals with a third dose on R_t_ by considering five scenarios which were combinations of vaccination, previous infection and contact rates that span the range of circumstances in many countries in late 2021. The five scenarios (with fraction vaccinated and previously infected shown for 1 September 2021) and rationale were (Table S7):

1. US/D (United States with distancing): R_0_ = 3.0, 54.6.% vaccinated, 58.4% previously infected: approximated fraction of US population previously infected with contact rates similar to summer-fall 2021
2. CA/D (California, USA, with distancing): R_0_ = 3.0, 58.7% vaccinated, 51.1% previously infected: approximated some populations with higher vaccination and lower fraction infected than scenario (1) (e.g. California)
3. NZ/ND (New Zealand, no distancing): R_0_ =7.0, 24.3% vaccinated, 0.2% previously infected: approximated countries with high vaccination rates that effectively suppressed transmission and had reduced contact rates when cases were detected (e.g. New Zealand, Australia, Hong Kong, etc.)
4. US/ND (United States, no distancing): R_0_ = 7.0, 54.6% vaccinated, 58.4% previously infected: a scenario to compare to scenario (1) to determine if boosting could limit transmission if behavior returned to pre-pandemic levels
5. US/ND-100 (United States, no distancing): R0 = 7.0, 100% vaccinated, 58.4% previously infected: a hypothetical optimistic scenario to compare to scenario (1) to determine if vaccination with or without boosting could limit transmission if behavior returned to pre-pandemic levels

We estimated the fraction of the population that was susceptible using data on the fraction of the population that had been vaccinated or infected or both. We first split each population into three groups based on vaccination status: fraction unvaccinated, f_U_, fraction vaccinated, f_V_, with two doses of either BNT162b2 or mRNA-1273, and fraction boosted with a third dose of the same vaccine used for the primary series, f_B_ (f_U_+f_V_+f_B_=1). We calculated the fraction of people in the USA receiving BNT162b2 and mRNA-1273 from vaccination records ^49^. We estimated the timing of infections using deconvolution of deaths as described above and calculated the number of infections by dividing deaths by an estimate of the infection fatality ratio (IFR) across of 2020-21 of 0.003 ^30^; the results were not sensitive to moderate variation in this estimate. We also examined estimating infections using cases and an infection/case ratio of 4.0 ^46^ which produced a lower estimate of total infections, and the reduction in R_t_ with third dose boosting and deaths averted were very similar to the death-deconvolution and IFR method. We accounted for reinfections by using daily infection-derived protection estimates for susceptibility and infectiousness produced by our model assuming neutralizing antibody titers had waned 90 days on average and using the NATR_var_ of the dominant circulating variant in the United States each day. We estimated the fraction of unvaccinated or vaccinated individuals that had been previously infected using the ratios of cases in these two groups over time in the USA ^50^ (Figure S5). We used vaccination and infection data to calculate the fraction of people previously infected among unvaccinated, f_PU_, and vaccinated, f_PV_ individuals (Table S7). We used rates of waning of antibody titers and data on the timing of vaccination and infection from the USA and New Zealand ^49^ for US and New Zealand scenarios, respectively, to estimate the distribution of NATR_tot_ in populations for the four time points in autumn 2021. We used the distributions of NATR_tot_ to estimate the relative susceptibility, 1-VE_S_, (Figure 1A) and relative infectiousness, 1-VE_I_, (Figure 1B) for the population in each of four groups: unvaccinated and previously infected (VE_SP_ and VE_IP_), vaccinated twice and no previous infection (VE_SV_ and VE_IV_), vaccinated three times or boosted (VE_SB_ and VE_IB_), and vaccinated and infected or hybrid immunity (VE_SH_ and VE_IH_), all with appropriate adjustments for waning antibody titers (see below). We used these estimates of VE to calculate R_t_ for populations composed of different fractions of five groups: never infected unvaccinated (1-f_PU_)f_U_, previously infected unvaccinated f_P_f_U_, previously uninfected vaccinated with two doses (1-f_PV_)f_V_, previously infected and vaccinated with two doses f_PV_f_V_, and boosted with a third dose, regardless of previous infection, f_B_:

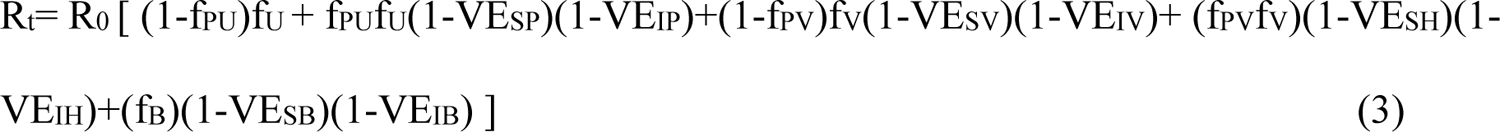

Note that in equation (3) the VE values for each population are integrated across the distribution of NATR_tot_ values for that group, and the distribution of NATR_tot_ values in each group reflects the timing of vaccination and infection on a daily timestep.

We used the scenario estimates of the fraction of population with different types of immunity (with timing matched to vaccination and infection and waning as described above) for susceptibility and infectiousness to estimate values of R_t_ under each scenario at the four time points in autumn 2021 (Sept. 1, Oct. 1, Nov. 1, and Dec. 1; Table S7). We calculated 95% CIs for predicted values of R_t_ that incorporated uncertainty in the relationship between time since vaccination or infection and NATR_istatus_, uncertainty in the relationships between neutralizing antibody titers and VE_S_ and VE_I_, as well as uncertainty in NATR_vac_ and NATR_var_ (Figures 1 and S2).

### The impact of third doses on infections and deaths caused by the Delta variant

We estimated the timing of the infections that gave rise to observed deaths and total infections in the United States in autumn 2021 using deconvolution as described above. We used SARS-CoV-2 sequencing data ^30,48^ to estimate the daily fraction of all infections that were caused by the Delta variant. We estimated the reduction in Delta infections if third-dose boosters were given to all fully vaccinated individuals at four time points in autumn 2021. We assumed the additional doses would take full effect simultaneously on Sept. 1, Oct. 1, Nov. 1, or Dec. 1. We estimated the number of Delta infections that would have occurred if third doses had been deployed by multiplying the number of estimated infections on each day by the reduction in the daily R_t_ due to third doses (e.g. 0.812 if R_t_ was reduced by 18.8%), raised to the power of the number of generations of infections (generation time 3.8d ^51^) since Sept 1 (or Oct. 1, Nov. 1 or Dec. 1). The calculated reduction in R_t_ was the difference between the number of actual third-doses given, and the number of doses deployed in the scenario. We estimated the number of deaths that would have been averted in each scenario by re-convoluting the reduced infections, multiplied by the IFR, using the delays described previously. We used uncertainty associated with our R_t_ estimates to calculate 95% CIs in the number of deaths averted. We rounded estimates and 95% CIs for infections and deaths to the nearest 100,000 and 100, respectively, to avoid giving a false sense of precision.

We used the estimates of VE_D_, the ratio of deaths in the unvaccinated compared to the vaccinated ^50^ (Figure S6), and the fractions of people who were unvaccinated, had received two doses, and had received a third-dose booster to estimate the number of deaths that would have been averted due to the direct protection against death if third-doses boosters were given to all fully vaccinated individuals at the same four time points. We repeated the calculation for the scenario where those same booster doses were instead used to provide two doses to unvaccinated individuals. We compared the number of deaths averted due to the reductions in transmission following third-dose boosting and the direct protection against death provided to individuals in these scenarios.

## Data availability

All data used in the paper are contained in the supplemental material.

R Code and data files to replicate the figures and analyses of this paper are available: https://github.com/marmkilpatrick/Vaccine-boosters

Upon publication, R Code will be uploaded to Zenodo and data files will be archived with Dryad.

## Acknowledgements

We received funding from the National Science Foundation (DEB-1911853, 1115895, and 1717498) and the California Department of Health.

## Author Contributors

BJG and AMK conceived the study, performed the analyses and wrote the paper.

