## Supplemental materials for "Deaths averted by vaccination due to reduced transmission can exceed those from direct protection of vaccinated individuals for SARS-CoV-2"

### Supplemental Results Text

We estimated the effect of boosting doubly-vaccinated individuals with a third dose of mRNA vaccines on $R_t$ for five scenarios, including the one presented in the main text, that had varying vaccination and infection rates and contact rates (Figure S2; see Methods for additional details). In countries or populations that had prevented most transmission (e.g. New Zealand) resulting in a very low cumulative fraction previously infected (~1%), and where vaccination was only partly underway (24.3% of the population) and where there was no social distancing (ND), resulting contact rates being at pre-pandemic levels, ($R_0$ = 7), boosting with a third dose would have reduced $R_t$ by only 5.3% from 5.68 to 5.38 and would have been insufficient to reduce $R_t$ below 1 to prevent a surge (Figure S2: Scenario NZ/ND, blue line). Similarly if contact rates in the US had returned to pre-pandemic levels in September, 2021, without social distancing ($R_0$ = 7), and with the observed vaccine coverage (53.2%) and infection history (67.4%), then boosting all doubly vaccinated individuals would have reduced $R_t$ by the same amount as the scenario in the main text,18.4%, since the change in VE would be identical, but $R_t$ would decrease from 2.57 to 2.10 (Figure S2: Scenario US/ND, purple line) rather than 1.17 to 0.96. This clearly would not have been sufficient to stop a huge surge in transmission (Figure S2: Scenario US/ND, purple line).

If the US population had originally achieved 100% vaccination coverage, but assuming the timing of vaccinations remained the same, and 67.4% had been previously infected, waning of vaccine and infection-derived immunity by late September 2021 would have led to a sizeable surge without boosting assuming pre-pandemic contact rates ($R_0$ = 7) (Figure S2: Scenario US/ND-100, left end of green line: $R_t$=1.14). However, boosting only 17.9% of this fully (100%)

vaccinated population would have reduced $R_t$ below 1 and thus could prevent a surge in cases (Figure S2 Scenario US/ND-100, green line crosses the $R_t = 1$ line at 17.9%). Boosting the entire population with a third dose would have reduced $R_t$ by 69.8% from 1.14 to 0.34 which would have essentially stopped transmission (Figure S2: Scenario US/ND-100, right end of green line: $R_t$=0.34).

**Supplemental Tables and Figures**

**Table S1. Statistics for the model fitting relationship between neutralizing antibody titers and VE given by the equation** $VE = 1 - \frac{1}{1+e^{-(c_0+c_1 log_2(NATR_{tot}))}}$.

| Endpoint | Coefficient | Estimate | SE | Z value | p-value |
|---|---|---|---|---|---|
| Susceptibility | $c_0$ | -1.066698 | 0.029608 | -36.027 | < 2.2e-16 |
| | $c_1$ | -0.410926 | 0.021249 | -19.339 | < 2.2e-16 |
| Infectiousness | $c_0$ | 0.174254 | 0.057717 | 3.0191 | 0.002535 |
| | $c_1$ | -0.506766 | 0.047208 | -10.7348 | < 2.2e-16 |

**Table S2. Analysis of relative waning rates of neutralizing antibodies relative to peak (Figure 2B) for two vaccines (BNT162b2 and mRNA-1273) and infection-derived immunity. The best fitting model by AIC included variation in slopes between vaccines and differences in asymptote between the two vaccines and infection-derived immunity: $log_2$(Antibody titer) = ($c_0$+$c_1$*Infection)*$e^{(c2*Day+c3*Day*mRNA\text{-}1273)}$-$c_0$+$c_1$*Infection, where BNT162b2 was the reference level. $c_0$ is the asymptote for long periods after vaccination for both vaccines. $c_1$ is the difference in asymptotes between infections and vaccines. We did not fit a separate asymptote for mRNA-1273 due to limited data > 6 months for waning in this vaccine. Hybrid waning rates have been shown to be similar to waning rates following two-dose vaccination [1].**

| Coefficient | Estimate | SE | t-value | P-value |
|---|---|---|---|---|
| $c_0$ (asymptote: time = ∞, BNT162b2/mRNA-1273) | 3.27 | 0.34 | 9.51 | <0.0001 |
| $c_1$ (asymptote: time = ∞, infection - $c_0$) | 1.55 | 0.33 | 4.76 | 0.0003 |
| $c_2$ (slope: BNT162b2/infection) | -0.012 | 0.0027 | -4.31 | 0.0006 |
| $c_3$ (slope: mRNA-1273 – $c_2$) | 0.043 | 0.0019 | 2.30 | 0.0364 |

**Table S3. Estimated vaccine effectiveness for two vaccines for susceptibility ($VE_S$) and infectiousness ($VE_I$) after waning and after a third vaccine dose using the relationship between neutralizing antibody titers and VE in Figures 1 and 2.**

| Vaccine, Endpoint | VE waned (95% CI) | VE boosted | Ratio |
|---|---|---|---|
| BNT162b2, $VE_S$ | 46.8% (35.4% - 56.9%) | 85.9% (80.4% - 90.1%) | 1.8 |
| BNT162b2, $VE_I$ | 16.3% (10.0% - 24.1%) | 67.6% (56.6% - 77.1%) | 4.1 |
| mRNA-1273, $VE_S$ | 54.3% (42.6% - 71.9%) | 84.5% (74.9% - 90.9%) | 1.6 |
| mRNA-1273, $VE_I$ | 22.0% (13.7% - 41.7%) | of 64.7% (46.7% - 79.3%) | 2.9 |

**Table S4. Effective reproductive number, fraction vaccinated, fraction infected, and a fraction of the population in four subpopulations in Figure 3 and Figure S2 for each time-location scenario.**

| Scenario | Month | $R_0$ | Frac vacc. | Frac inf. | Frac vacc. and inf. ($f_{PV}$ * $f_V$) | Frac. vacc. and not inf. ((1- $f_{PV}$) * $f_V$) | Frac. not vacc. and inf. ($f_{PU}$ * $f_U$) | Frac. fully susc. ((1-$f_{PU}$) * $f_U$) |
|---|---|---|---|---|---|---|---|---|
| US/D | Sept. 1 | 3.2 | 0.53 | 0.67 | 0.32 | 0.21 | 0.35 | 0.12 |
| | Oct. 1 | 3.2 | 0.56 | 0.73 | 0.36 | 0.20 | 0.37 | 0.07 |
| | Nov. 1 | 3.2 | 0.58 | 0.76 | 0.38 | 0.19 | 0.38 | 0.04 |
| | Dec. 1 | 3.2 | 0.59 | 0.81 | 0.41 | 0.18 | 0.40 | 0.01 |
| CA/D | Sept. 1 | 3.2 | 0.59 | 0.60 | 0.33 | 0.26 | 0.27 | 0.15 |
| | Oct. 1 | 3.2 | 0.62 | 0.62 | 0.35 | 0.26 | 0.27 | 0.12 |
| | Nov. 1 | 3.2 | 0.63 | 0.64 | 0.38 | 0.26 | 0.27 | 0.10 |
| | Dec. 1 | 3.2 | 0.65 | 0.66 | 0.40 | 0.26 | 0.26 | 0.09 |
| NZ/ND | Sept. 1 | 7 | 0.24 | 0.002 | 0.00 | 0.24 | 0.001 | 0.76 |
| | Oct. 1 | 7 | 0.38 | 0.002 | 0.00 | 0.38 | 0.001 | 0.62 |
| | Nov. 1 | 7 | 0.62 | 0.002 | 0.001 | 0.62 | 0.00 | 0.38 |
| | Dec. 1 | 7 | 0.71 | 0.003 | 0.002 | 0.71 | 0.001 | 0.29 |
| US/ND | Sept. 1 | 7 | 0.53 | 0.67 | .32 | 0.21 | 0.35 | 0.12 |

| | | | | | | | | |
|---|---|---|---|---|---|---|---|---|
| | Oct. 1 | 7 | 0.56 | 0.73 | 0.36 | 0.20 | 0.37 | 0.07 |
| | Nov. 1 | 7 | 0.58 | 0.76 | 0.38 | 0.19 | 0.38 | 0.04 |
| | Dec. 1 | 7 | 0.59 | 0.81 | 0.41 | 0.18 | 0.40 | 0.01 |
| US/ND-100 | Sept. 1 | 7 | 1.00 | 0.67 | 0.67 | 0.33 | 0 | 0 |
| | Oct. 1 | 7 | 1.00 | 0.73[26] | 0.73 | 0.27 | 0 | 0 |
| | Nov. 1 | 7 | 1.00 | 0.76 | 0.76 | 0.24 | 0 | 0 |
| | Dec. 1 | 7 | 1.00 | 0.81 | 0.81 | 0.19 | 0 | 0 |

**Table S5. Estimated infections and deaths averted (95% CI) given third-dose boosting that takes full effect on September 1, October 1, November 1, or December 1.**

| **Month** | **Infections averted (95% CI)** | **Deaths averted (95% CI)** |
|---|---|---|
| September 2021 | 37,147,423 (34,530,588 – 38,860,029) | 111,442 (103,592 – 116,580) |
| October 2021 | 24,503,688 (23,045,328 – 25,328,986) | 73,511 (69,136 – 75,987) |
| November 2021 | 12,124,437 (11,014,685 – 12,788,302) | 36,373 (33,044 – 38,365) |
| December 2021 | 1,984,987 (1,671,856 – 2,209,045) | 5,955 (5,016 – 6,627) |

**Table S6. Delay distributions used for deconvoluting deaths to estimate the timing of infections.**

| **Delay** | **Distribution** |
|---|---|
| Infection to shedding | Gamma (shape = 5.983, scale = 1.455) |
| Shedding to symptom onset | Weibull (shape = 0.294, scale = 0.14) |
| Symptom onset to hospitalization | Gamma (shape = 5.078, scale = 0.765) |
| Hospitalization to death | Gamma (shape = 2.1, scale = 6.524) |

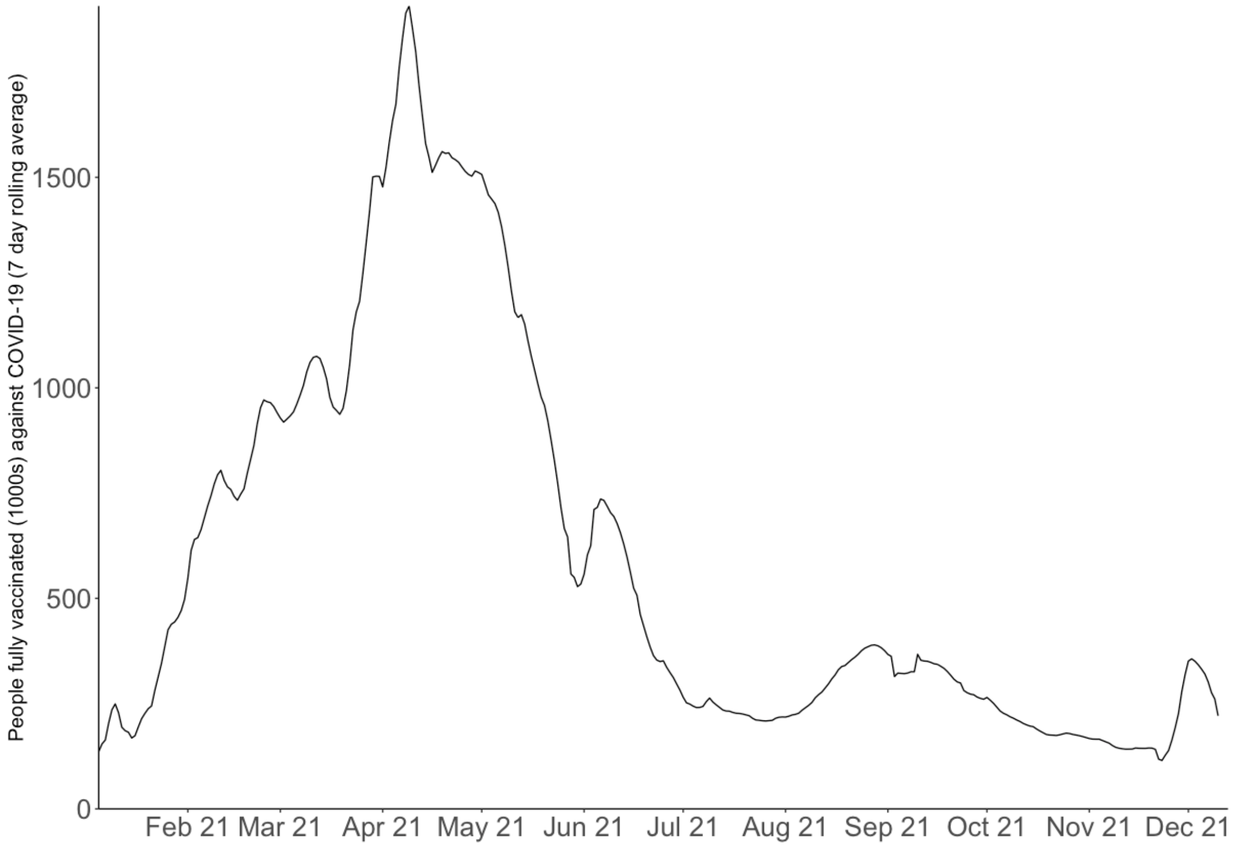


**Figure S1. Number of people fully vaccinated in the USA[2].**

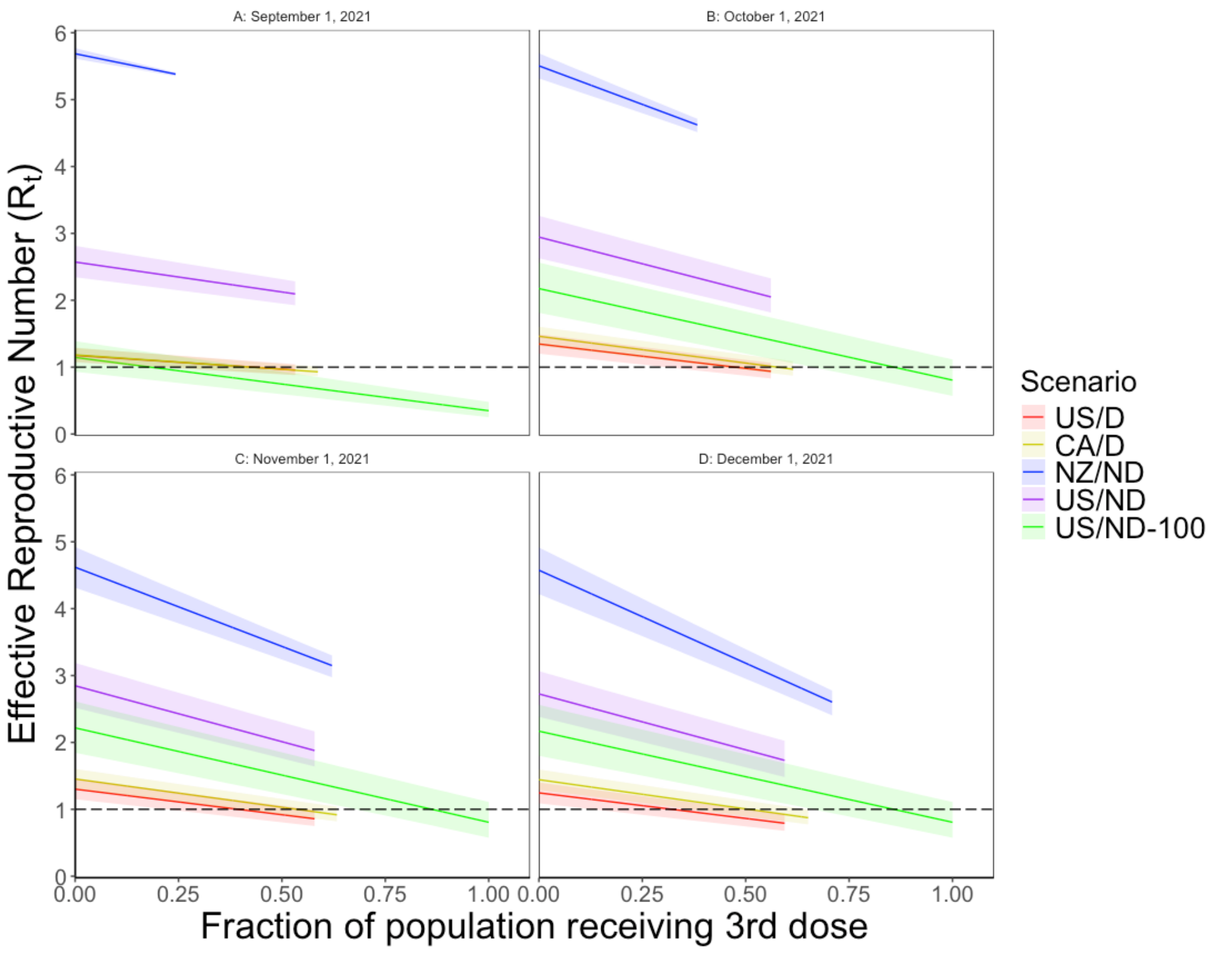


**Figure S2. Lines and 95% CIs show $R_t$ for five scenarios (see legend) which vary in location (CA – California, NZ – New Zealand, US – United States) with location-specific vaccination and infection as of September 1, October 1, November 1, and December 1, 2021, distancing (D) or no distancing (ND) which determine contact rates and $R_0$ values. See Table S5 for time-location-specific R0, infection rates, and vaccination rates.**

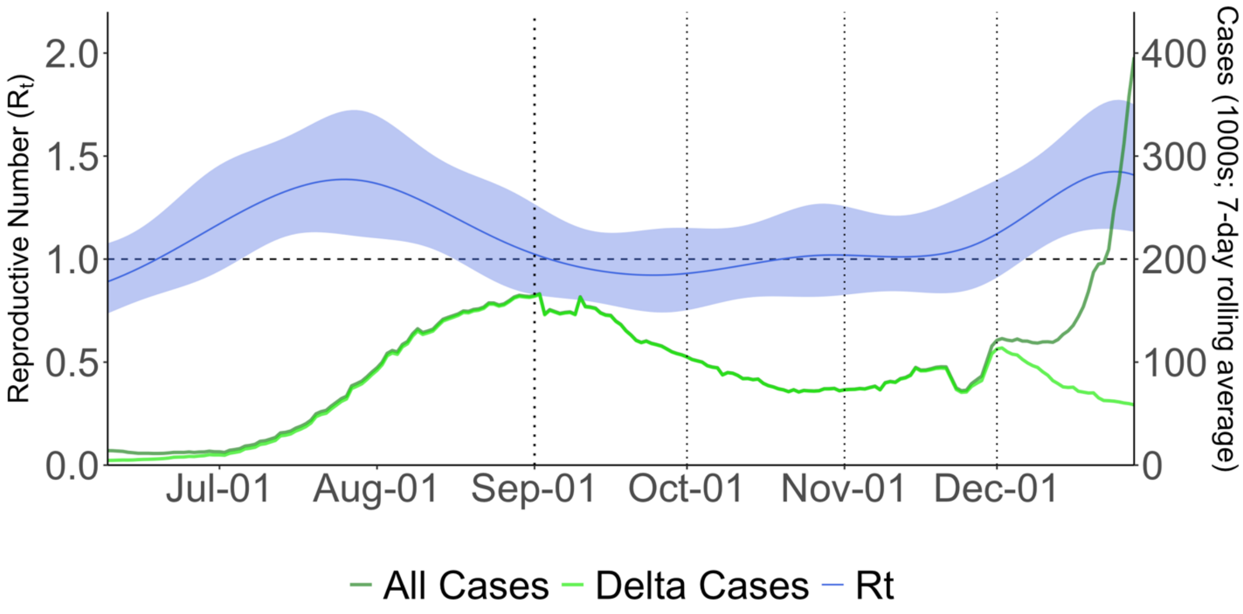


**Figure S3. Reproductive number, $R_t$ (blue line and ribbon) and daily COVID-19 (all and delta-specific) cases for the USA during the period when the Delta variant was prevalent, July 2021-January 2022. Dotted lines show four separate dates considered for third doses to take full effect.**

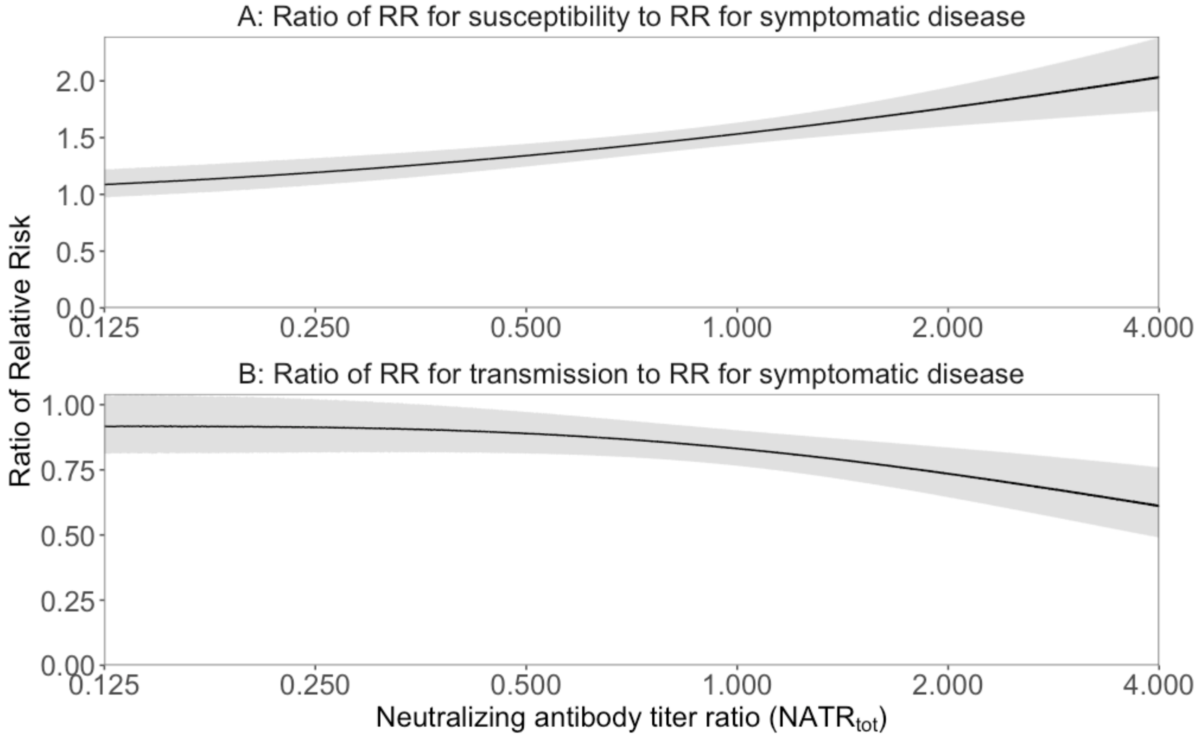


**Figure S4. Lines and 95% CI for the (A) ratio of relative risk (1 – VE) for susceptibility from Figure 1A to relative risk for symptomatic disease using estimates from [3] and (B) ratio of relative risk for transmission from Figure 1C to relative risk for symptomatic disease [3] plotted against neutralizing antibody titer ratios ($NATR_{tot}$).**

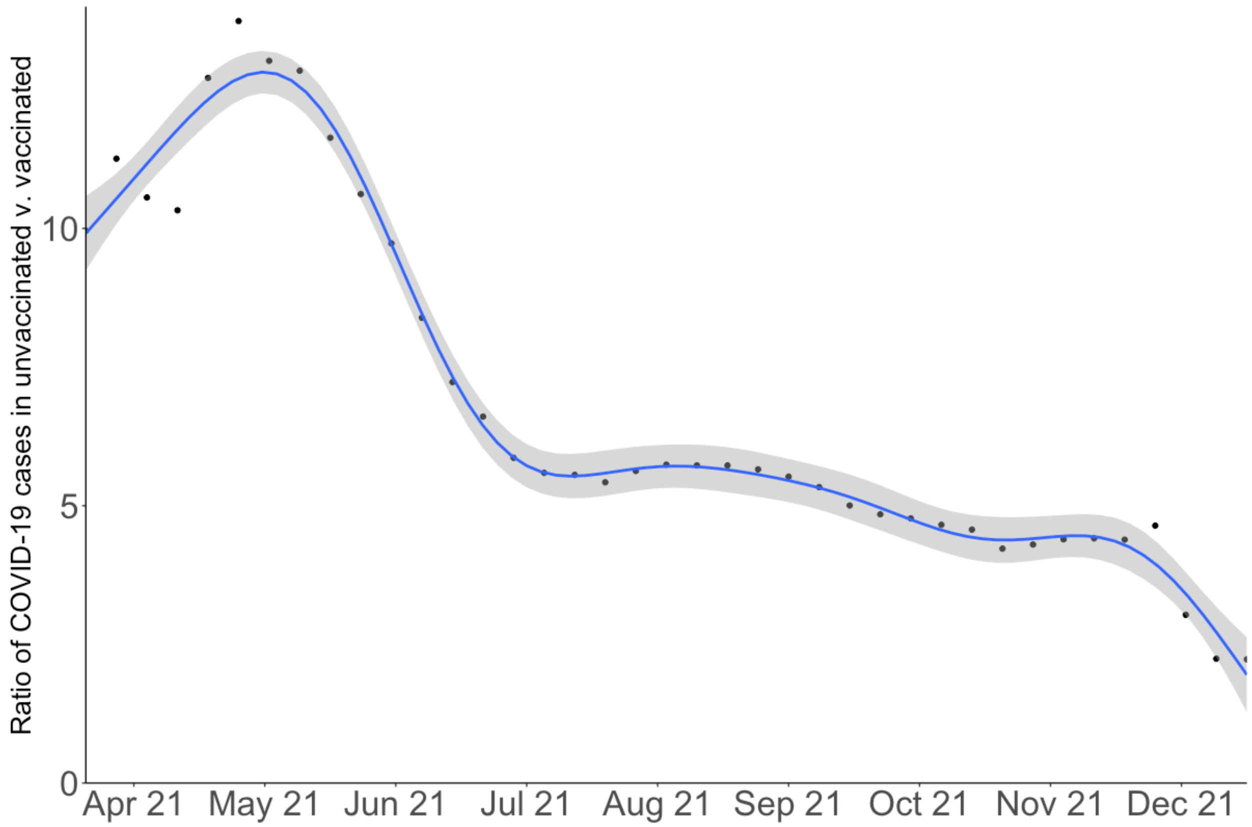


**Figure S5. Ratio of COVID-19 cases in unvaccinated individuals relative to vaccinated individuals in the USA in 2021 based on data from CDC (CDC, 2021d). Points show weekly values, and the line and ribbon shows a generalized additive model fit and 95% CI.**
